# Changes in RT-PCR-positive SARS-CoV-2 rates in adults and children according to the epidemic stages

**DOI:** 10.1101/2020.05.18.20098863

**Authors:** Corinne Levy, Romain Basmaci, Philippe Bensaid, Cécile Bost Bru, Edeline Coinde, Emmanuelle Dessioux, Cécile Fournial, Jean Gashignard, Hervé Haas, Véronique Hentgen, Frédéric Huet, Muriel Lalande, Alain Martinot, Charlotte Pons, Anne Sophie Romain, Nicoleta Ursulescu, François Vie Le Sage, Josette Raymond, Stéphane Béchet, Julie Toubiana, Robert Cohen

## Abstract

**Aim:** To describe the trends of RT-PCR positive SARS-CoV-2 rates in children and adults according to the time of COVID-19 epidemic.

**Methods:** In this prospective multicenter study involving 45 pediatric units, we collected the results of nasopharyngeal swabs in France from March 2, 2020 to April 26, 2020.

**Results:** During the study period, 52,588 RT-PCR tests for SARS-CoV-2 were performed, 6,490 in children and 46,098 in adults. The risk ratio of RT-PCR positive SARS-CoV-2 tests for adults compared to children was 3.5 (95% CI [3.2;3.9]) for the whole study period. These rates varied according to the time of the epidemic and were higher at the peak. The lower rates of positive test in children persisted during the surveillance period but varied according to the time in the epidemic.

**Conclusion:** The rate of positive RT-PCR positive SARS-CoV-2 tests for children was always less than that for adults but vary according to the epidemic stage.

## Introduction

France has been markedly affected by COVID-19, with more than 83,000 hospitalized cases at the epidemic peak.^1^ The mortality and morbidity of this virus is highly variable among age groups.^2, 3^ In France and in other countries, the number of confirmed pediatric cases is relatively low, and they account for less than 1% of hospitalized cases and deaths.^3-6^ In France, the strategy of closing schools and the lockdown started on March 17, 2020, but little information is available about the transmission between children and adults.^4, 7^

We aimed to describe the trends of RT-PCR–positive SARS-CoV-2 rates in children and adults as compared with the profile of the national epidemic curve of new COVID-19 cases in France.^6^

## Methods

With the Association Clinique et Thérapeutique Infantile du Val de Marne (ACTIV) and Groupe de Pathologie Infectieuse Pédiatrique (GPIP) network research units, we conducted a French prospective multicenter study involving 45 hospitals, pediatric wards, emergency units, and virology laboratories which performed RT-PCR analysis for SARS-CoV-2 from March 2, 2020 to April 26, 2020. The strategy of closing schools and the lockdown decided by the French government for the whole country started on March 17 and finished on May 11, 2020. Here, we collected aggregate data of RT-PCR–positive SARS-CoV-2 rates for each hospital: the proportion of children with positive results out of children tested and the proportion of adults with positive results out of adults tested. During the study period, the patients visiting emergency departments or who were hospitalized were selected for RT-PCR SARS-CoV-2 testing if they had severe disease (adults and children), and/or they were contact with a confirmed COVID-19 case, and/or they were healthcare workers (adults) with symptoms. Twice a week, each clinical investigator from each participating ward was contacted to obtain information on SARS-CoV-2–positive tests.

Risk ratio and 95% confidence intervals (95% CIs) of RT-PCR–positive SARS-CoV-2 tests for adults compared to children were calculated using Stata SE v13.1 (Statacorp, College Station, TX).

## Results

During the study period, 52,588 RT-PCR tests for SARS-CoV-2 were performed, 6,490 in children (12.3%) and 46,098 in adults (87.7%). The cumulative rate of positive tests for children was 5.9%, (n=382), 3.5-fold less than that for adults, 20.3%, (n=9,346).

The figure 1 shows the trends of positive testing in children and adults in France and in the Paris area, one of the most affected regions, as well as the overall national trend in new Covid-19 cases.

**Figure 1.**
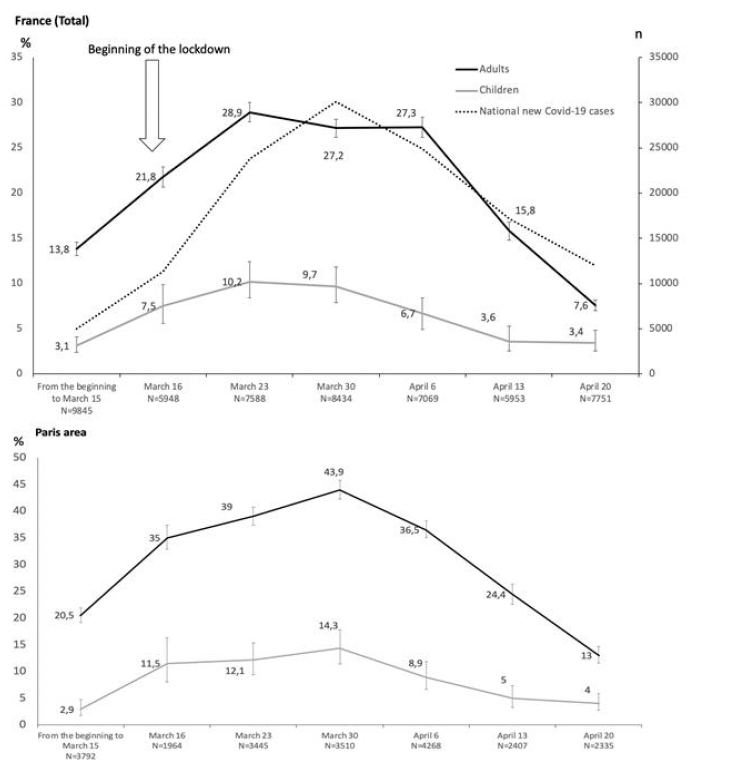
Evolution of RT-PCR–positive SARS-CoV-2 rates in France and in Paris area in children and adults as compared with new COVID-19 cases reported by the National Health Institute. ^6^

In France, from the beginning of the epidemic until March 15, only 3.1% (n=53) of 1,690 pediatric samples were positive, 4.5-fold less than for adults (13.8%, n=1,124 of 8,155 adult samples). At the peak of the national outbreak, on March 30, 9.7% (n=85) of 877 pediatric samples were positive, 2.8-fold lower than for adults, 27.2% (n=2054 of 7,557 adult samples). A rapid decrease was observed during the following weeks, with the lowest rate reported the week of April 20, 3.4% (n=33) of 960 pediatric samples were positive, 2.2-fold lower than for adults, 7.6% (n=514 of 6,791 adult samples).

In Paris area, the same trends were observed with marked differences between adults and children.

The risk ratio of RT-PCR–positive SARS-CoV-2 tests for adults compared to children was 3.5 (95% CI [3.2;3.9]) for the entire period of the study. The Figure 2 showed the evolution of this risk ratio in France and in the Paris area week per week during the study period. For Paris area, at the beginning of the epidemic, the risk ratio was 7.1 (95% CI [4.3; 11.7]), whereas it ranged from 3 and 4.9 during the following weeks.

**Figure 2.**
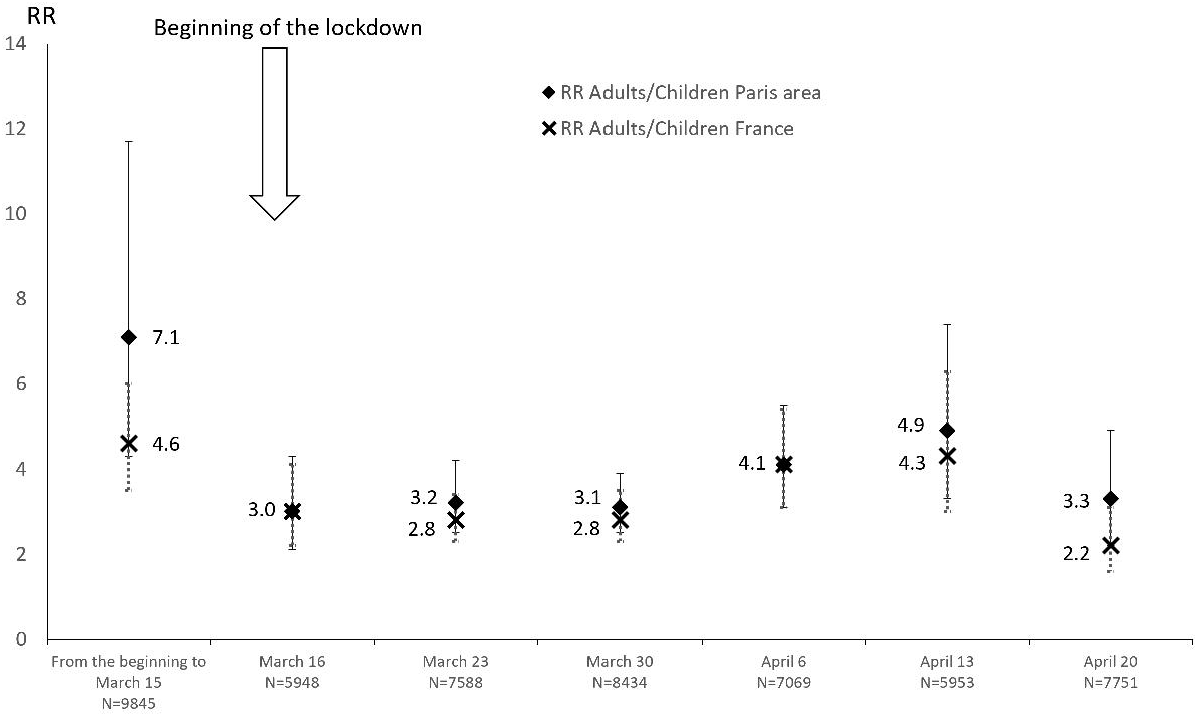
Evolution of the risk ratio of RT-PCR–positive SARS-CoV-2 tests for adults compared to children in France and in the Paris area during the study period.

## Discussion

Our results showed in a large cohort (n=52,588), that the risk ratio of RT-PCR–positive SARS-CoV-2 tests for adults compared to children was 3.5 for the entire period of the study. However, the dynamics of the curve for children followed that for adults and the National curve for new COVID-19.^1^ The difference in rates between children and adults persisted during the surveillance period but varied according to the time in the epidemic, the rate of positivity and the region. In the Paris area, the most affected region, we observed a high spread of the disease in children, reaching more than 14.3% positive tests at the epidemic peak. At the beginning of the epidemic, the risk ratio was 7.1 (95% CI [4.3;11.7]), whereas it ranged from 3 and 4.9 during the following weeks.

As expected, the major impact of the lockdown was observed about 15 days after it began, with a rapid decrease in SARS-CoV-2–positive rate. Of note, the more the rate in adults increased, the less the difference between the two populations (adults and children), which supports that the main way of transmission was from adults to children.^4, 5 7^ Indeed, in this study, as in many others, the rate of RT-PCR–positive SARS-CoV-2 was significantly lower for children than adults.^5, 8^Even if the viral load is comparable for children and adults, this fact added to the low rates of secondary cases at schools argue simultaneously for at least a modest role of children in the dynamic of the COVID-19 pandemic and the re-opening schools.^4, 7 9 10^

The slight increase of the risk ratio of RT-PCR–positive SARS-CoV-2 tests for adults compared to children just before the end of the epidemic is possibly explained by the occurrence of Kawasaki like syndromes or hyperinflammatory shock in children 2 to 4 weeks after the peak epidemic.^11, 12^

Our study has several limitations. Our rates of RT-PCR–positive SARS-CoV-2 must take into account that RT-PCR practices were heterogeneous and could have evolved depending on the guidelines and the availability of the tests for adults and children. Even if we cannot rule out that some cases could have escaped our surveillance in our centers and that we did not survey the whole country, our results are consistent with the dynamic of the COVID-19 pandemic in France.^6^ Finally, the viral load was not available in our study for children and adults.

We think that the surveillance of RT-PCR–positive tests in adults and children could be a simple and reliable tool to survey the epidemiology of SARS-CoV-2 infection, allowing to quickly detect any re-emergence of the disease.

## Data Availability

data are available upon reasonable request

## Acknowledgments

We thank all pediatricians and microbiologists who participated in the study:

Belgaid A, Belivier E, Bueno B, Brehin C, Canarelli J, Castain L, Caurier B, Chevret L, Claris O, Cohen L, Colonna de Cinarca V, De Barbentane MC, De Pontual L, De Rougemont A, Dolfi-Fiette H, Dutron S, Faye A, Fernandez F, Flatres C, Foulongne V, Gajdos V, Garraffo A, Guillotel E, Hau I, Jarlier V, Jeziorski E, Kuentz M, Labarthe F, Laisney N, Landraud L, Larrat S, Le Stradic C, Lina B, Loeile C, Mandelcwajg A, Marques C, Minodier P, Morand A, Morvan O, Ouldali N, Pantalone L, Peigne C, Pierre MH, Pinquier D, Raoult D, Rey A, Teissier R, Thach C, Varon E, Vignaud O.

## Contributors

CL, RC and VH conceived and designed the study. CL and RC were involved in writing, reviewing and coordinating the submission of the manuscript. RB, ASR, JT, PB, CBB, EC, ED, CF, JG, HH, FH, ML, AM, CP, NU, FLS, JR contributed to data collection from their respective paediatric centres and have reviewed the manuscript. SB have reviewed the manuscript.

## Patient consent for publication

Not required

## Ethics approval

The GFHGNP (Groupe Francophone d’Hépatologie et Nutrition Pédiatrique) Ethics Comittee, certifies that this study “Surveillance of SARS CoV2 positive PCR in France” performed by the GPIP ACTIV doesn’t required any ethical approval due to the absence of personal data.

